# Objectively measured physical activity among people with and without HIV in Uganda: associations with cardiovascular risk and coronary artery disease

**DOI:** 10.1101/2024.06.07.24308634

**Authors:** Chinonso C. Opara, Christine Horvat Davey, Cissy Kityo, Ellen Brinza, Rashidah Nazzindah, Marcio Summer Bittencourt, Vitor Oliveira, Allison R. Webel, Chris T. Longenecker

## Abstract

**Background:** Africa has a disproportionate burden of HIV-related cardiovascular disease. We aimed to describe physical activity in people living with HIV (PLHIV) and people without HIV (PWOH) in Uganda and characterize its relationship with the presence of computed tomography angiography-detected (CCTA) coronary artery disease (CAD).

**Methods:** We performed a cross-sectional analysis of the Ugandan Study of HIV Effects on the Myocardium and Atherosclerosis using Computed Tomography (mUTIMA-CT) cohort. From 2017-2019, physical activity in PLHIV and PWOH was assessed by accelerometry over seven days. Participants additionally underwent CCTA. Univariable and multivariable modified Poisson regression was used to analyze the relationship between physical activity and CAD presence.

**Results:** 168 participants were analyzed. The median (IQR) age was 57 (53-58) years old and 64% were female. Males had more moderate-to-vigorous physical activity per week [68 minutes (12-144) vs 15 minutes (0-50), *P*<0.001] and less light physical activity [788 minutes (497-1,202) vs [1,059 (730-1490), *P*=0.001] compared to females, but there was no difference by HIV status. After adjusting for age, which accounted for 10% of the variation in steps taken, and sex, no significant associations were found between physical activity and coronary plaque.

**Conclusion:** Objectively measured physical activity was low compared to guideline recommendations, with males being somewhat more active than females and without significant differences by HIV status. Physical activity was not associated with the presence of CAD independently of age and sex.

**CLINICAL PERSPECTIVE:** *What is new?:*

- For the first time, we describe objective physical activity patterns in people living with HIV (PLHIV) and people without HIV (PWOH) in the Ugandan context. Overall, few people met guideline recommendations for physical activity. Males had more moderate-to-vigorous physical activity and were more likely to meet guideline recommendations compared to females, without significant difference by HIV status.
- In contrast, females had more light physical activity compared to males. Although light physical activity appeared to have a stronger inverse relationship with coronary artery disease (CAD) in PLHIV compared to PWOH in stratified models, the relationship was significantly confounded by age and sex.

*What are the clinical implications?:*

- Larger studies are needed to further investigate the relationship between physical activity and CAD in Africa—including any modifying influence of sex or HIV status. In the meantime, our study suggests there is significant room to enhance physical activity in Uganda.
- Age informs the relationship between physical activity and CAD, and our study suggests efforts to emphasize exercise as the population ages may play a role in reducing CAD burden.

## INTRODUCTION

Africa is disproportionately impacted by HIV and HIV-related cardiovascular disease (CVD). Mirroring outcomes in higher-resourced countries, increased life-expectancy due to more widely available antiretroviral therapy (ART) has led to higher rates of age-related comorbidities, such as CVD, non-Autoimmune Deficiency Syndrome (AIDS) malignancies and liver disease^1^. Over the past two decades, the global burden of HIV-associated CVD has tripled and accounts for 2.6 million disability-adjusted life-years per year, with the highest impact in Africa and the Asia-Pacific areas^2^. Key risk factors for CVD in people living with HIV (PLHIV) include hypertension^3^, diabetes mellitus^4^, dyslipidemia^5,6^, obesity^3^, inflammation^6^, low nadir CD4+ count, and smoking^3^.

Physical activity is a key preventive factor for CVD. Physical activity and exercise training interventions improve cardiovascular health, decrease CVD risk, and mitigate morbidity and mortality in those with CVD^6^. For instance, eight weeks of moderate intensity exercise resulted in improved aerobic fitness, reduced blood pressure and increased CD4 cells for PLHIV in Nigeria^7^. Despite the proven benefits, the majority of PLHIV in Africa do not seem to engage in regular physical activity^8^. However, most prior research has relied on self-reported measures of physical activity, which has been shown to overestimate physical activity compared to gold-standard objective measures of physical activity, such as accelerometry^9^.

Atherosclerotic cardiovascular disease (ASCVD) risk can be assessed using risk prediction scores or objective measures of subclinical disease; however, risk prediction scores may not strongly correlate with subclinical disease in Africa. For example, research conducted in Uganda and Botswana demonstrated only modest correlations between pre-clinical atherosclerosis measures and the pooled cohort equation’s ASCVD risk score^3^. More research is needed to address non-traditional risk factors unique to this region, including infectious and environmental factors such as latent tuberculosis and air pollution^3^. In the meantime, enhancing physical activity and fitness is a low-cost approach to ASCVD risk reduction in Africa.

Our study was designed to compare objective measures of physical activity among PLHIV and a similar population of people without HIV (PWOH), which has not previously been described in Uganda. Secondly, we aimed to evaluate the relationship between physical activity metrics and ASCVD risk, coronary artery disease (CAD) presence by coronary computed tomography angiography (CCTA), and severity of CAD by coronary CTA. Lastly, we sought to assess whether HIV status and/or sex modify the relationship between physical activity and CAD.

## METHODS

### Participant Selection

The **<u>U</u>**gandan s**<u>T</u>**udy of H**I**V effects on the **<u>M</u>**yocardium and **<u>A</u>**therosclerosis using **C**omputed **T**omography (mUTIMA-CT), conducted in Kampala, Uganda, is an ongoing prospective cohort investigation examining the impact of HIV on the myocardium and atherosclerosis. In this phase of the study, PLHIV were age-(within 3 years) and sex-matched in a 1:1 ratio with PWOH controls. PLHIV were recruited from the Joint Clinical Research Centre (JCRC) in Lubowa, Uganda, which is near Kampala. PWOH were recruited from community or hospital-based internal medicine clinics in Kampala. To be eligible for inclusion, PLHIV must have been on ART for more than six months without any changes in regimen within 12 weeks prior to enrollment. All participants, regardless of HIV status, were ≥ 45 years old and had at least one cardiovascular disease (CVD) risk factor, such as hypertension, low high-density lipoprotein cholesterol, diabetes mellitus, smoking, or a family history of early CAD. Exclusion criteria were a history of known CVD, peripheral artery disease, ischemic stroke, uncontrolled chronic inflammatory conditions, pregnancy, use of chemotherapy or immunomodulating agents, or an estimated glomerular filtration rate (eGFR) less than 30 ml/minute.

The original cohort comprised of 100 PLHIV and 100 PWOH enrolled from April 2015 to May 2017. Previous publications have detailed the findings and methods of the baseline examination of the original cohort^10–13^. Follow-up examinations were conducted from 2017 to 2019, with total cohort size of 200 participants maintained by replacing those lost to follow-up with age- and sex-matched individuals. This follow-up consisted of 2 study visits separated by 7 days, during which physical activity metrics were collected via accelerometry. This current study is a cross-sectional analysis of those data among participants with accelerometry data duration of at least 10 hours per day on at least 4 days^14,15^, which is described as valid wear time.

The study protocol was approved by the University Hospitals Cleveland Medical Center Institutional Review Board, the JCRC Research Ethics Committee, and the Uganda National Council for Science and Technology, with all participants providing written informed consent prior to any study procedures.

### Physical Activity

Self-reported measures of physical activity were assessed using the International Physical Activity Questionnaire – Short Form (IPAQ-SF). This was developed as a tool for international monitoring of physical activity and sedentary time^16^. The IPAQ-SF contains a series of 7 questions, requiring participant recall of light, moderate, and vigorous physical activity time, as well as sedentary time over the preceding 7 days. The amount of light, moderate, and vigorous physical activity per day was multiplied by the number of days performing those activities to get the amount of activity per week. The amount of time spent sitting per day was multiplied by 7 to obtain the amount of sedentary time per week.

Objective measures of physical activity were assessed by accelerometry. Participants were instructed to wear an Actigraph®wGT3X+-BT monitor (Actigraph, LLC, Pensacola, FL) for 7 days on their non-dominant hip. Physical activity metrics of interest were steps per day, light physical activity time per week, moderate-to-vigorous physical activity time per week, and sedentary time per week. The number of steps per day was derived by dividing the number of total steps recorded by the number of valid wear days. Other physical activity metrics were derived by dividing the amount of activity time by the number of valid days and multiplying by 7. Data was sampled at 30 Hz, using 60-second epochs and the normal filter. Bouted physical activity was used to measure physical activity accumulation over 10 minutes or more. Different intensities of physical activity were defined by adult cut-points for tri-axial accelerometers^17^. In this validation, moderate-to-vigorous activity was defined as achieving at least 3 metabolic equivalents and was predicted by achieving at least 2,960 triaxial counts per minute. Valid wear time was determined by data recorded for at least 10 hours per day for at least 4 days. Accelerometry data was analyzed using Actilife software.

### Atherosclerotic Cardiovascular Disease Risk

10-year ASCVD risk was quantified using the pooled-cohort equation^18^. The following variables were utilized: age, sex, and smoking status were obtained by self-report; diabetes was defined as a chart confirmed diagnosis or on medication for diabetes; hypertension treatment was defined as receiving medication to lower blood pressure. Systolic and diastolic blood pressure were obtained at the initial visit by trained study staff. Total cholesterol, high density lipoprotein, and calculated low-density lipoprotein concentrations were measured by the JCRC clinical lab from fasting blood drawn at the initial visit. Notably, we chose to use the “other race” term, since the African American term has not been validated in the East African population. Further, the nature of atherosclerotic cardiovascular disease risk may be different in the East African population compared to the population from which the pooled-cohort equation was derived^10^. Elevated ASCVD risk was defined as 10-year ASCVD risk ≥5%, which is the threshold to consider use of statin therapy to reduce ASCVD risk^18^.

### Coronary Artery Disease

At visit 2, all participants underwent coronary computed tomography angiography (CCTA) on a 128-slice Siemens Somatom scanner at Nsambya St. Francis Hospital in Kampala. Participants were ineligible to have intravenous contrast for CCTA if their estimated GFR was not greater than 60 mL/min/1.73m^2^. Data acquisition was additionally limited for some participants by other technical reasons (e.g., inability to adequately lower the heart rate with betablockers) (Figure 1). The acquisition and image analysis protocols were developed in alignment with the Society of Cardiovascular Computed Tomography Guidelines^19^. Participants were given 100 mg of oral metoprolol two hours prior to the scan. They were additionally given another 50 mg dose 30 minutes prior to the scan if the heart rate was elevated above 60 beats per minute.

**Figure 1.**
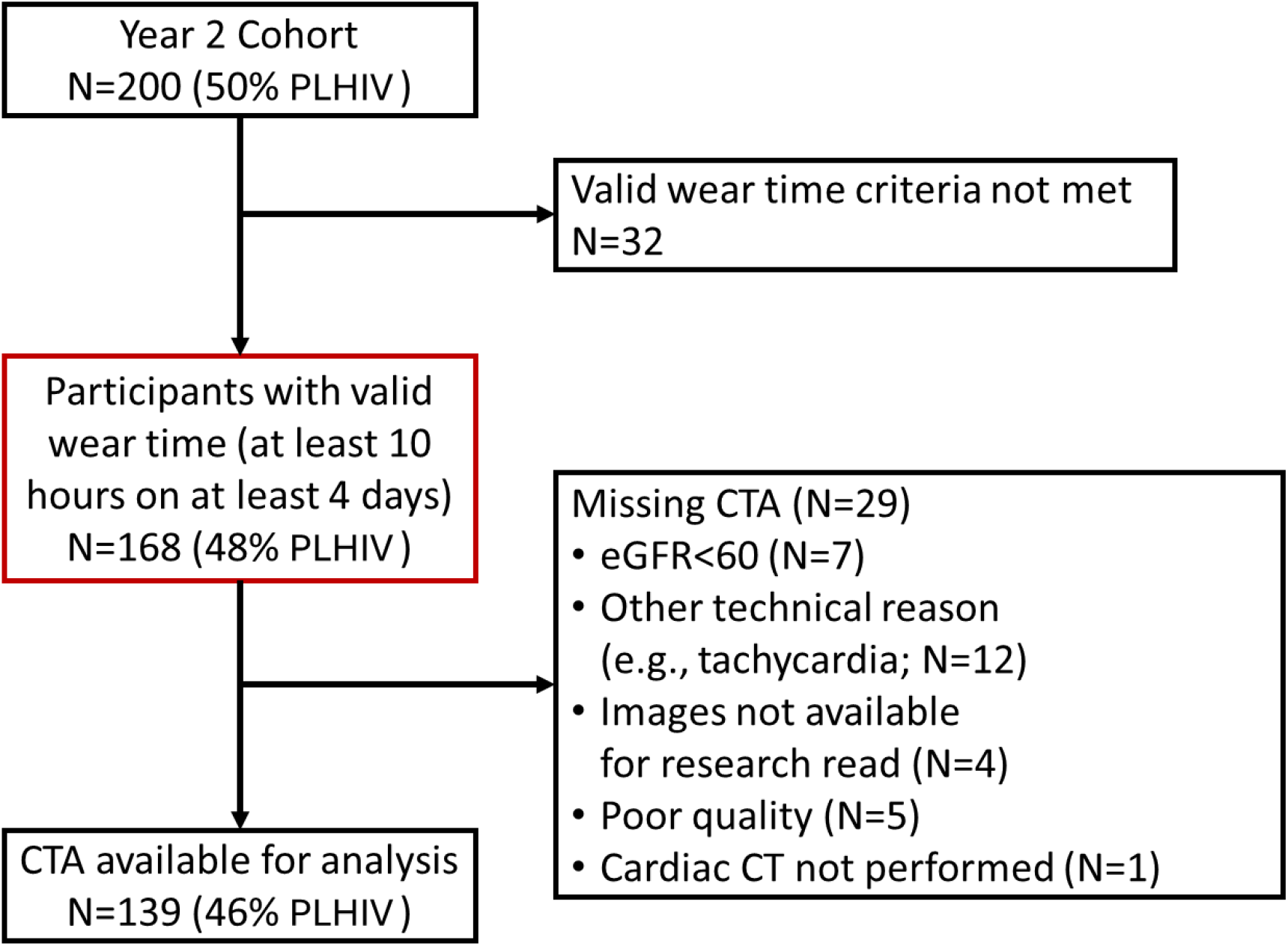
Derivation of the study population from the mUTIMA-CT cohort. Definitions: PLHIV, people living with HIV; CTA, computed tomography angiography; eGFR, estimated glomerular filtration rate.

The CT scans were first read by a local radiologist for clinically significant findings and were subsequently read offline in batch by a single expert reader (MSB) for research. Scans that were of poor technical quality were excluded from analysis. Segment involvement score (SIS) was defined as the total number of disease segments.

Segment severity score (SSS) was calculated using a luminal obstruction weight for that segment (x1 if <25% obstruction, x2 if 25–50%, x3 if 50–70%, x4 if 70–99% and x5 if totally occluded), giving a maximum possible SSS of 90 for an 18-segment model. The presence of CAD was defined as SIS>0 and more severe CAD was defined as an SSS greater than the median among those with CAD.

### Statistical Analysis

Demographic and clinical characteristics were described by frequency (percent) for categorical variables and median (interquartile range) for continuous variables, stratified by HIV status and sex. Physical activity measures were described in a similar manner. Spearman’s correlation coefficient was used to evaluate the relationship between self-reported and gold-standard objective measures of physical activity. Bland-Altman plots were used to assess the degree of agreement between the two measures of physical activity. The objective measures were subtracted from the self-reported measures to obtain measurement differences and the limits of agreement were defined as the mean difference ± 1.96 times the standard deviation of the differences. Spearman’s correlation was additionally used to assess the relationship between age and objective measures of physical activity. Additionally, the Wilcoxon rank-sum test was used to compare physical activity metrics in their continuous version across binary subgroups of HIV status and sex. Chi-square tests were used to compare the proportion of those achieving at least 150 minutes of moderate-to-vigorous exercise per week and those achieving at least ten thousand steps per day by HIV status and sex.

Unadjusted and adjusted Poisson regression models with robust standard errors were used to estimate prevalence ratios to analyze the relationship between binary measures of physical activity, by median cut points, with elevated ASCVD risk, presence of CAD among those with available CTA data, and presence of more severe CAD among those with CAD. Binary measures of physical activity were used to meet the linearity assumption of log transformation of the outcomes of interest with their physical activity predictors. Median cut points were used to allow for sufficient sample size on both sides of the selected cut points. Covariates – age and sex – were chosen a priori to build the adjustment models. HIV status and sex interaction terms, along with likelihood ratio tests, were used individually to assess for effect modification in the relationship between binary objective physical activity metrics and the presence of CAD. A schematic of the data analysis approach may be found in Supplementary Figure 1.

R version 4.3.1 was used for analysis: *P*<0.05 was considered statistically significant.

## RESULTS

### Study cohort

The cohort began with 200 participants (Figure 1). The overall cohort for this analysis included 168 participants with valid accelerometry wear time, 48% of whom were PLHIV. 139 participants among those with valid accelerometry wear time had coronary angiography available for analysis (46% PLHIV). The most common reasons for missing coronary angiography were tachycardia and low eGFR.

### Baseline characteristics of study participants

The median (IQR) age of the overall cohort was 57 (53, 62) years old and 64% were female (Table 1). The median age was similar across HIV status and sex. 84% of PLHIV had viral load suppression, and males were more likely to have HIV viral load suppression than females (94% vs 77%). Overall, smoking rates were low (2.4%) and no females reported smoking. The median systolic blood pressure was higher in males than in females [155 mmHg (139-171) vs 146 (131-162)]. Males had higher ASCVD 10-year risk than females [13% (8-24) vs 6% (3-10)]. Males had lower body mass index than females [25 (23-28) vs 31 (27-34)].

**Table 1.**
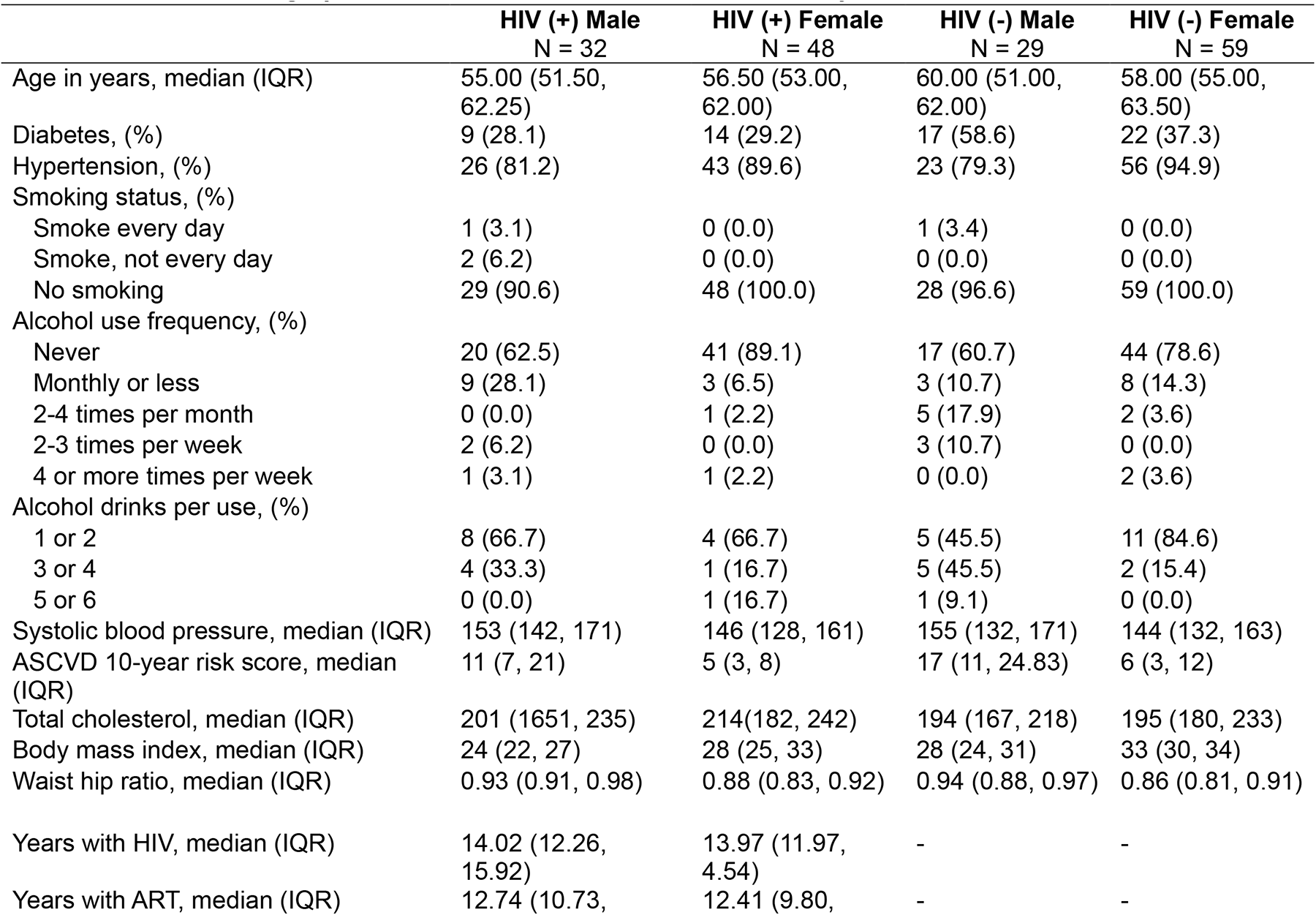
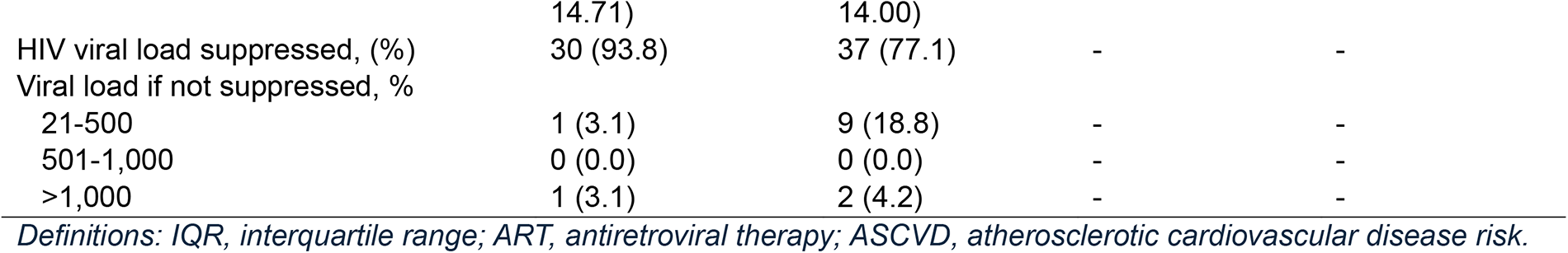
Baseline demographic and clinical characteristics, stratified by HIV status and sex.

**Table 2.** Objectively measured physical activity stratified by (a) HIV status and (b) sex.

|  |  |  |  |
| --- | --- | --- | --- |
| **(a). Stratified by HIV status** |  |  |  |
|  | **PLHIV, n=80** | **PWOH, n=88** | **P-value** |
| Valid wear days, median (IQR) | 6.00 (6.00, 7.00) | 6.00 (5.75, 7.00) | 0.993 |
| Total step counts per day, median (IQR) | 5859.05 (4308.05, 7735.14) | 5814.08 (4027.48, 7556.35) | 0.574 |
| Light activity time per week in minutes, median (IQR) | 949.08 (666.87, 1369.08) | 934.00 (649.03, 1342.00) | 0.881 |
| Moderate-to-vigorous activity time per week in minutes, median (IQR) | 18.38 (0.00, 95.88) | 35.50 (0.00, 74.31) | 0.889 |
| Sedentary time per week in minutes, median (IQR) | 1234.50 (951.56, 1806.58) | 1,301.00 (883.31, 1959.13) | 0.906 |
| Achieved 150 minutes moderate-to-vigorous activity or more per week, (%) | 11 (13.8) | 7 (8.0) | 0.335 |
| Achieved 10 thousand steps or more per day, (%) | 9 (11.2) | 6 (6.8) | 0.462 |
| **(b). Stratified by sex** |  |  |  |
|  | **Male, n=61** | **Female, n=107** | **P-value** |
| Valid wear days, median (IQR) | 6.00 (6.00, 7.00) | 6.00 (6.00, 7.00) | 0.757 |
| Total step counts per day, median (IQR) | 6534.00 (5073.00, 9177.50) | 5382.33 (3807.58, 7316.42) | 0.005 |
| Light activity time per week in minutes, median (IQR) | 788.00 (497.00, 1201.67) | 1059.00 (730.40, 1490.42) | 0.001 |
| Moderate-to-vigorous activity time per week in minutes, median (IQR) | 68.00 (11.67, 143.50) | 15.17 (0.00, 49.50) | <0.001 |
| Sedentary time per week in minutes, median (IQR) | 1320.67 (1079.17, 1850.33) | 1203.00 (873.25, 1916.83) | 0.303 |
| Achieved 150 minutes moderate-to-vigorous activity or more per week, (%) | 13 (21.3) | 5 (4.7) | 0.002 |
| Achieved 10 thousand steps or more per day, (%) | 10 (16.4) | 5 (4.7) | 0.023 |

*Definitions: IQR, interquartile range; PLHIV, people living with HIV; PWOH, people without HIV*

**Table 3.** Unadjusted and adjusted associations of objective physical activity metrics with ASCVD 10-year risk ≥ 5%, n=168.

|  | Unadjusted |  | Adjusted |  |
| --- | --- | --- | --- | --- |
|  | PR (95% CI) | P-value | PR | P-value |
| Total step counts per day ≥ 5.9 thousand | 0.954 (0.770-1.181) | 0.663 | 1.042 (0.850-1.277) | 0.694 |
| Light activity time per week ≥ 15.7 hours | 0.807 (0.649-1.002) | 0.053 | 0.991 (0.821-1.195) | 0.922 |
| Moderate-to-vigorous activity time per week ≥ 0.4 hours | 0.909 (0.734-1.126) | 0.383 | 0.934 (0.770-1.134) | 0.491 |
| Sedentary time per week ≥ 21 hours | 1.049 (.847-1.299) | 0.663 | 0.864 (0.714-1.045) | 0.131 |

*Definitions: ASCVD, atherosclerotic cardiovascular disease risk; PR, prevalence ratio; CI, confidence interval. Adjusted for age and sex.*

**Table 4.**
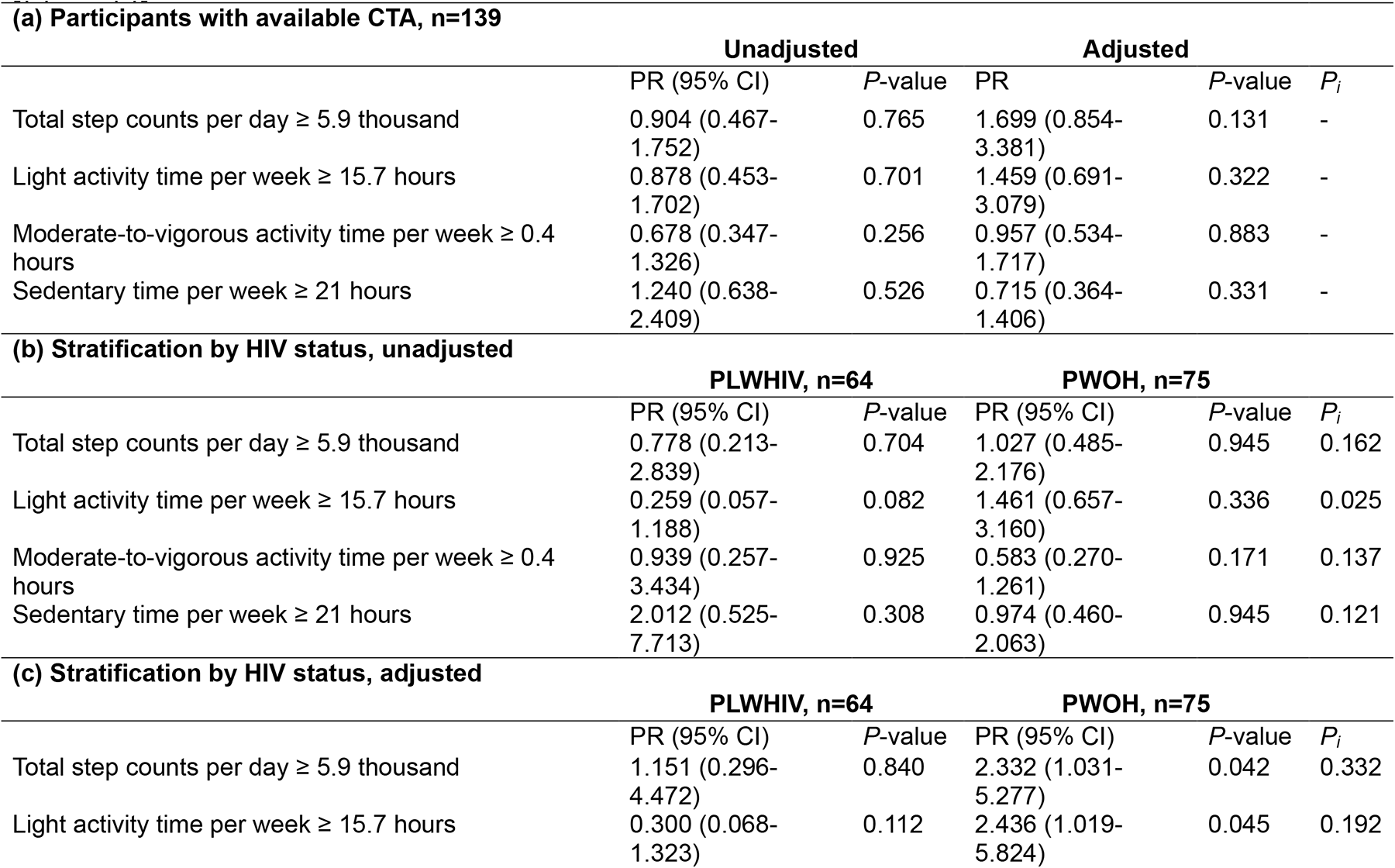
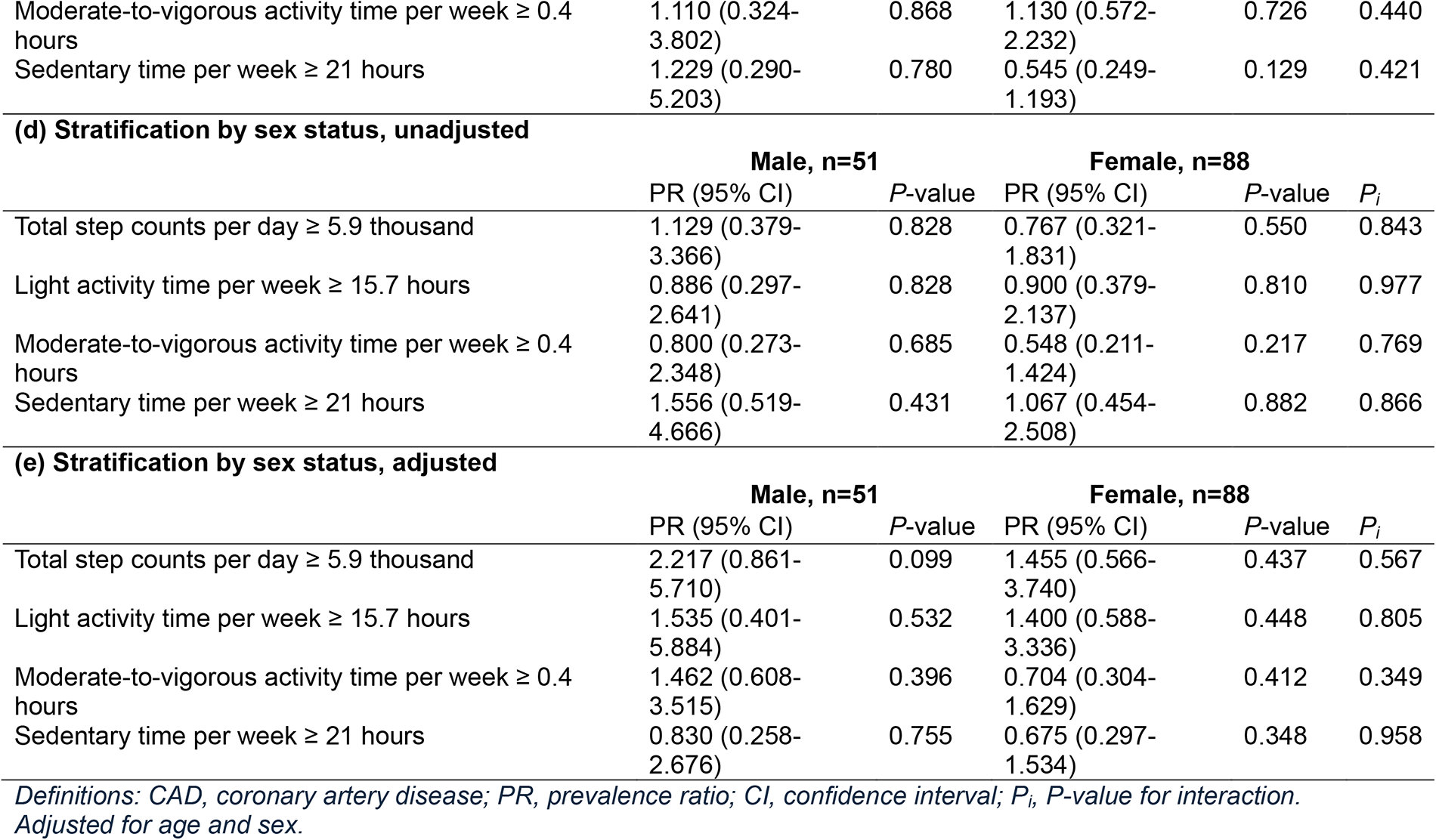
Unadjusted and adjusted associations of objective physical activity metrics with presence of CTA-detected CAD for (a) the overall cohort with available CTA, stratified by HIV status [(b) and (c)] and by sex [(d) and (e)]

**Table 5.** Unadjusted and adjusted associations of objective physical activity metrics with more severe CTA-detected CAD, n=28.

|  | Unadjusted |  | Adjusted |  |
| --- | --- | --- | --- | --- |
|  | PR (95% CI) | P-value | PR | P-value |
| Total step counts per day ≥ 5.9 thousand | 0.429 (0.138-1.329) | 0.142 | 0.480 (0.128-1.797) | 0.276 |
| Light activity time per week ≥ 15.7 hours | 0.429 (0.138-1.329) | 0.142 | 0.464 (0.138-1.559) | 0.214 |
| Moderate-to-vigorous activity time per week ≥ 0.4 hours | 0.571 (0.185-1.763) | 0.330 | 0.624 (0.199-1.955) | 0.418 |
| Sedentary time per week ≥ 21 hours | 2.022 (0.653-6.262) | 0.222 | 1.676 (0.464-6.051) | 0.430 |

*Definitions: CAD, coronary artery disease; PR, prevalence ratio; CI, confidence interval. Adjusted for age and sex.*

### Relationship between self-reported and objective measures of physical activity

There was no statistically significant correlation between self-reported measures of physical activity and objective gold-standard measures of physical activity (ρ=0.06, *P*=0.529 for sedentary time; ρ=-0.04, *P*=0.703 for light activity time; ρ=-0.16, *P*=0.172 for moderate-to-vigorous activity time; Supplementary Figure 2). Additionally, participants overestimated their moderate-to-vigorous activity and underestimated their light activity; and these errors in estimation were enhanced as self-reported or objectively measured physical activity increased. Objective measures of physical activity were used for the remainder of the analysis.

### Objective physical activity outcomes of study participants

The median (IQR) accelerometry valid wear days for the total cohort was 6 (6-7) days during the 7 days in which objective measures of physical activity were collected. The median step counts per day was higher for males than for females [6,534 steps (5,073-9,178) vs 5,382 steps (3,808-7,316), *P=* 0.005], and males were more likely to achieve at least 10,000 steps per day than females (16% vs 5%, *P*=0.023). Males had more moderate-to-vigorous physical activity per week [68 minutes (12-144) vs 15 minutes (0-50), *P*<0.001] than females and a greater proportion of men achieved at least 150 minutes of moderate-to-vigorous physical activity per week than females (21% vs 5%, *P*=0.002). Males had less light physical activity per week [788 minutes (497-1,202) vs [1,059 (730-1490), *P*=0.001] than females. There was no significant difference in objective physical activity outcomes when comparing PLHIV and POWH.

There was a statistically significant positive correlation between age and sedentary time (Spearman’s correlation coefficient, ρ=0.22, *P*=0.004) and a statistically significant negative correlation between age and moderate-to-vigorous physical activity (ρ=-0.25, *P*=0.001), and steps per day (ρ=-0.31, *P*<0.001) (Supplementary Figure 3). The negative correlation between age and light physical activity was not statistically significant (ρ=-0.12, *P*=0.113).

### Association of objective physical activity outcomes and atherosclerotic cardiovascular disease risk and coronary artery disease

Prior to adjusting for age and sex, participants who achieved at least 15.7 hours of light physical activity per week (the median in the total cohort) had non-statistically significant 19% lower prevalence of elevated ASCVD risk compared to those who had less than 15.7 hours of light physical activity per week (unadjusted prevalence ratio (PR) 0.807, 95% CI 0.649-1.002, *P*=0.053). This association was weakened after adjustment for age and sex. Prior to adjustment, there was a general non-statistically significant association of decreased risk of elevated ASCVD risk with more physical activity and increased risk of elevated ASCVD risk with more sedentary time for the overall cohort.

For the total cohort, no statistically significant association was found between physical activity metrics and presence of CCTA-detected CAD prior to and after adjustment. Prior to adjustment, there was a general non-statistically significant trend of lower risk of CTA-detected CAD with more physical activity and greater risk of CAD with greater sedentary time for the overall cohort.

After stratifying by HIV status and prior to adjustment, there was a trend of lower risk of CT-detected CAD in PLHIV with more light physical activity compared to PWOH [unadjusted PR 0.259, 95% CI 0.057-1.188, *P*=0.082 vs unadjusted PR 1.461, 95% CI 0.675-3.160, *P*=0.336] with a statistically significant interaction *P*-value of 0.025. After adjustment, the interaction was no longer statistically significant.

Similarly, there was no significant interaction of sex on the relationship between physical activity metrics and the presence of CTA-detected CAD prior to and after adjustment.

Prior to and after adjustment, there was a general non-statistically significant trend of lower risk of more severe CTA-detected CAD with more physical activity and higher risk of more severe CTA-detected CAD with more sedentary time.

## DISCUSSION

Self-reported measures of physical activity, such as IPAQ-SF have been described as a cost-effective method to assess physical activity^20^. In this study, we collected and compared self-reported and objective measures of physical activity by way of the IPAQ-SF and accelerometry, respectively. We demonstrated self-reported moderate-to-vigorous activity was overestimated while self-reported light activity was underestimated, and we did not find a statistically significant correlation between self-reported and objective physical activity metrics. These results were consistent with other investigations seeking to validate IPAQ-SF^9^, and highlights the potential role of objective measurement instruments in physical activity studies.

This is the first study to describe objective measures of physical activity among PLHIV and PWOH living in Uganda. While we did not find a significant difference in physical activity patterns between well-matched PLHIV and PWOH, we found significant differences between males and females. On average, males had 21% more steps per day and spent almost five times as much time engaging in moderate-to-vigorous physical activity. On the other hand, females spent 34% more time in light physical activity. These results are generally in line with traditional gender roles among males and females observed in Uganda^13^. Importantly, objectively measured physical activity was low in this population in urban Uganda, as only 11% of participants overall achieved moderate-to-vigorous physical activity time of at least 150 minutes, with just 5% of females achieving this target. Several studies have demonstrated the all-cause and cardiovascular mortality benefit from achieving this level of physical activity^21–23^, and this target has been recommended by several international cardiovascular organizations for the mitigation of ASCVD^24–27^.

A large volume of population-level evidence supports the reduction in ASCVD events with increased physical activity^28,21–23^, and this benefit may be mediated by an inverse relationship between physical activity and sub-clinical cardiovascular disease^29^. Despite this, we did not find a significant association between the varied levels of physical activity and inactivity with elevated ASCVD risk after adjusting for age and sex. Similarly, we did not find a significant association between physical activity metrics with presence of CTA-detected CAD or severity of CAD with multivariable analyses. However, our univariable analyses demonstrate an overall non-statistically significant trend of lower risk of elevated ASCVD risk, CTA-detected CAD, and CAD severity with higher levels of physical activity metrics for the overall cohort, with an opposite relationship with increased sedentary time. This trend was maintained in the multivariable analysis for CAD severity. Additionally, we demonstrated that increased light physical activity had a non-statistically significant association with lower prevalence of elevated ASCVD risk, with a 19% prevalence reduction for the total cohort. We also demonstrated a significant moderating effect of HIV, such that the inverse relationship between light physical activity and CAD was stronger among PLHIV compared to PWOH. While larger studies are needed to further characterize this finding, it does suggest that even light levels of physical activity may help reduce the burden of HIV-related cardiovascular disease.

We further assessed the correlation of age with physical activity metrics. We found that age was positively correlated with sedentary time. We demonstrated that age was negatively correlated with moderate-to-vigorous physical activity time and steps per day and explained as much as 10% of the variation in steps per day. We further demonstrated a trend of negative correlation between age and light physical activity time. These correlations help inform the relationship between physical activity metrics and ASCVD risk, CAD, and presence of more severe CAD within the Ugandan context. Given the trends of physical activity benefits seen in this study, it may indicate a role for encouraging lifelong exercise, especially as one ages. This further attracts attention to the potential need to promote physical activity in Africa, especially as it continues to industrialize.

### Strengths and Limitations

This work contains several important strengths including the use of gold-standard objective measures of physical activity and atherosclerotic CAD. We have shown that self-reported physical activity is not a good measure of objectively measured physical activity, arguing for the use of objective measures whenever feasible. Our prior research has also shown that ASCVD risk scores do not correlate well with objectively measured subclinical atherosclerosis in Uganda, and thus risk scores should not be used as a surrogate marker of subclinical ASCVD^10^.

However, our study also has limitations. First, it is possible that our sample size was not sufficient to detect significant associations between physical activity metrics and the outcomes described. Secondly, this was a cross-sectional analysis where measures of physical activity were assessed for a short period of 7 days. Although wearing the physical activity monitor during 7 days with a minimum of 4 days of valid data is recommended^30^, this did not capture the majority of lifetime physical activity habits that would inform the relationship with ASCVD risk, CAD, and severity of CAD. Thirdly, it is possible some participants may have altered their physical activity habits with the awareness of monitoring. Additionally, as this was an observational study, the univariable trends and associations seen cannot be construed as causal. Lastly, this was a single-center study based near the commercial capital of Uganda and the results might not be generalizable to those living in more rural areas.

## CONCLUSION

In this unique study from Africa, objectively measured physical activity was low in Urban Uganda compared to guideline recommendations, with males being somewhat more active than females and without significant differences by HIV status. Physical activity was not statistically significantly associated with the presence of CAD independently of age and sex. However, these hypothesis generating findings require further research with larger studies.

## Data Availability

All relevant data supporting the findings of this study are included within the manuscript and its supplementary information.

---

**Supplementary Figure 1.**
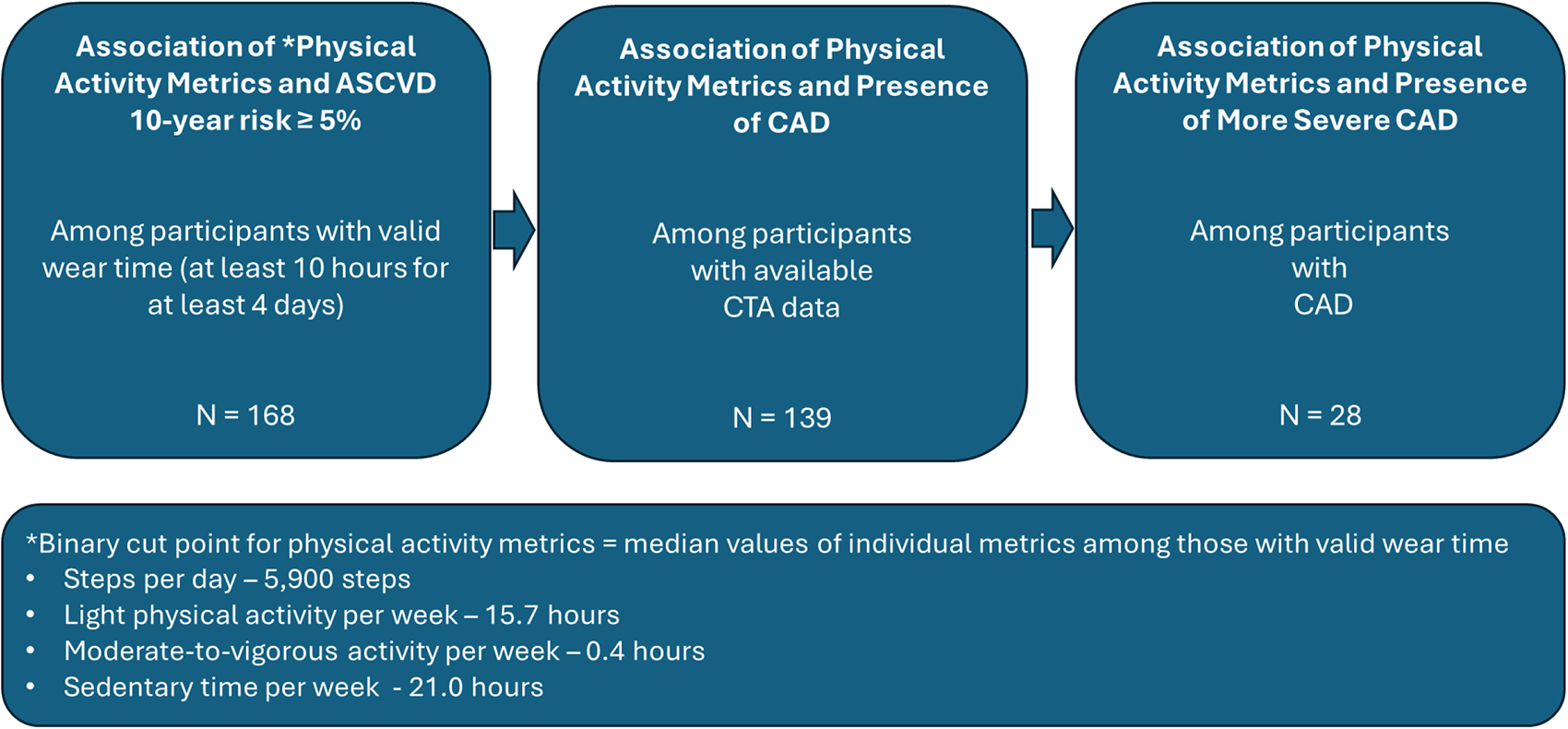
Depicts flow of data analysis among study participants. Objective physical activity metrics were used for analysis. Definitions: ASCVD, atherosclerotic cardiovascular disease risk; CAD, coronary artery disease.

**Supplementary Figure 2.**
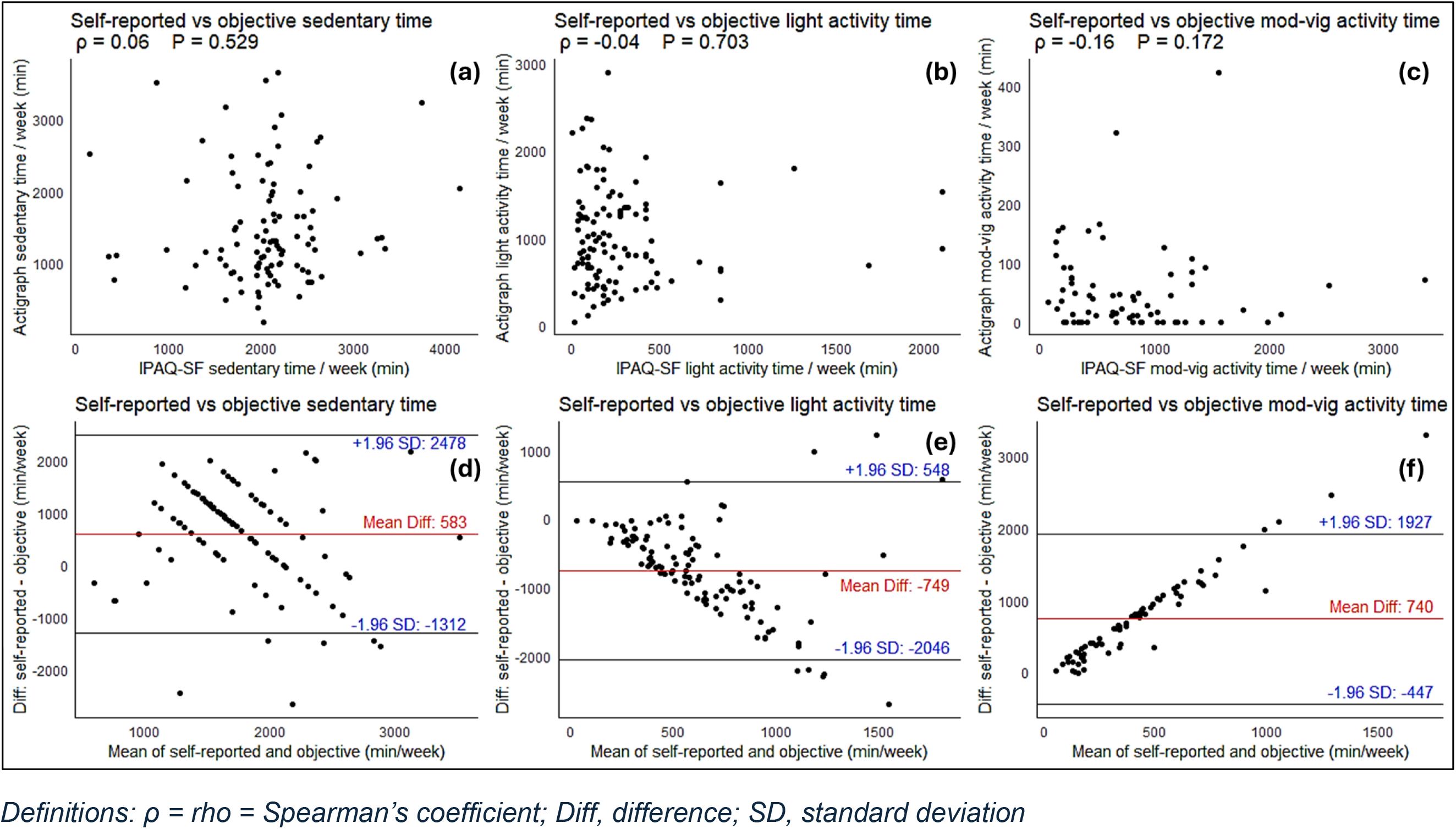
Relationship between self-reported and objective measures of physical activity. (a), (b), and (c) demonstrate scatter plots and Spearman’s correlation for self-reported vs objectively measured sedentary, light activity, and moderate-to-vigorous activity time, respectively. (d), (e), and (f) demonstrate Bland-Altman plots discerning the degree of agreement between self-reported vs objectively measured sedentary, light activity, and moderate-to-vigorous activity time.

**Supplementary Figure 3.**
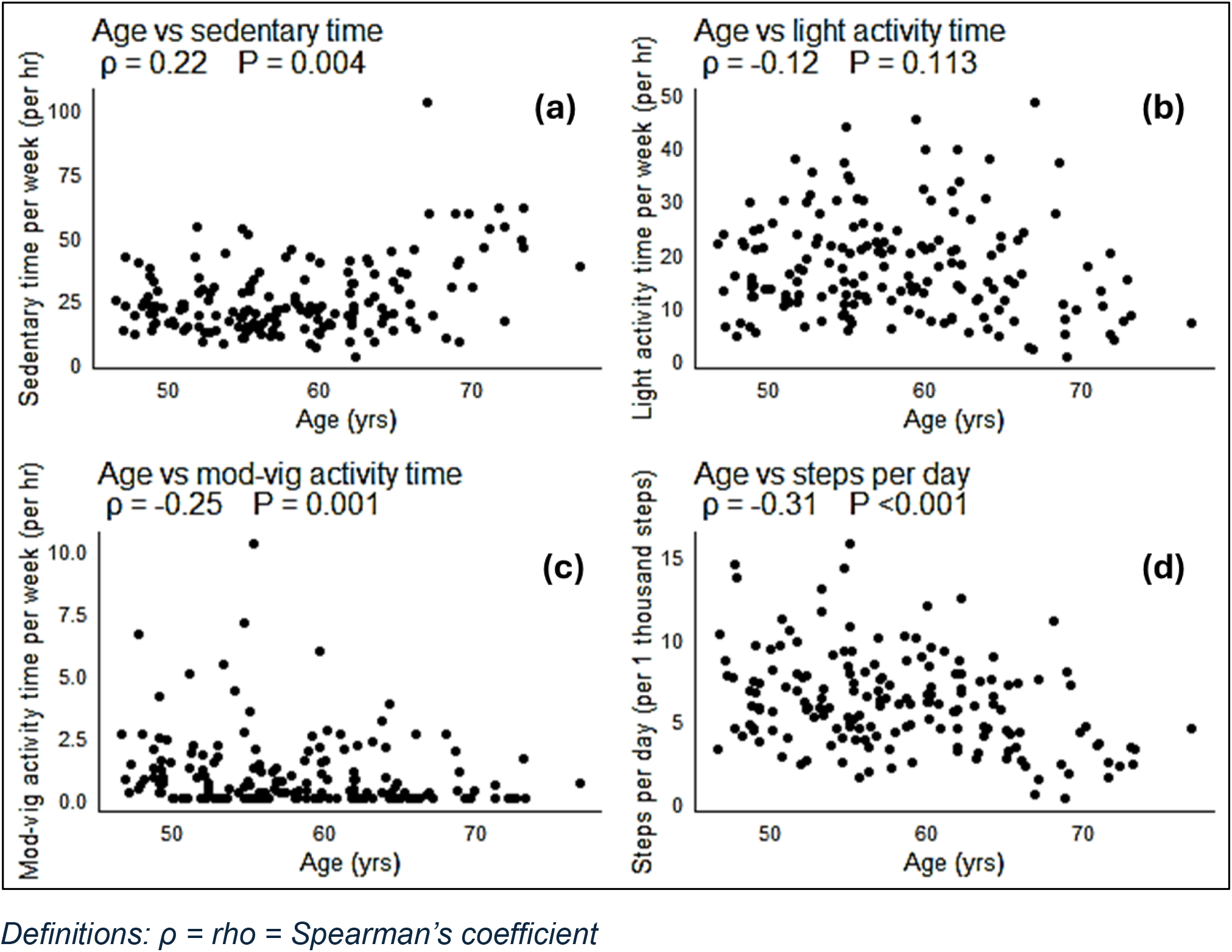
Correlation of age with objective measures of (a) sedentary time, (b) light activity time, (c) moderate-to-vigorous activity time, and (d) steps per day.

